# Cognitive Effects of *Toxoplasma* and CMV Infections: A Cross-Sectional Study of 557 Young Adults Considering Modulation by Sex and Rh Factor

**DOI:** 10.1101/2024.03.04.24303698

**Authors:** Jaroslav Flegr, Veronika Chvátalová, Lenka Příplatová, Petr Tureček, Petr Kodym, Blanka Šebánková, Šárka Kaňková

## Abstract

One-third of humanity harbors a lifelong infection with *Toxoplasma gondii*. This parasite undergoes sexual reproduction in cats and asexual reproduction in any warm-blooded intermediate hosts. The cycle progresses as cats ingest these hosts, containing the parasite’s tissue cysts. Such infections can alter behaviors in both animals and humans, potentially increasing predation risk by felines—usually seen as parasite-induced manipulations. This study aims to delineate toxoplasmosis’s effects on cognitive abilities and compare these to the effects of human cytomegalovirus (CMV), which also infects the brain but is not spread through predation. We evaluated the cognitive performance of 557 students, who had been examined for *Toxoplasma* and CMV infections, using intelligence, memory, and psychomotor tests. Results indicated cognitive impairments in seropositive individuals for both pathogens, with variations in cognitive impact related to sex and Rh factor. Specifically, *Toxoplasma* was associated with lower IQ in men, whereas CMV predominantly with worse women’s memory and reaction speeds. Analysis of antibody concentrations hinted that certain *Toxoplasma*-associated cognitive detriments may wane (impaired intelligence) or worsen (impaired reaction times) over time following infection. The findings imply that cognitive impairments from both neurotropic pathogens are likely due to pathological changes in the brain rather than direct manipulative actions by the parasites.

## 1. Introduction

*Toxoplasma gondii* is a protozoan parasite that undergoes sexual reproduction in its definitive hosts, any species of a cat. A broad spectrum of homoiotherm vertebrates, including humans, serve as intermediate hosts where *Toxoplasma* reproduces asexually. Approximately one-third of the global human population is infected with *Toxoplasma* [1]. Historically, latent *Toxoplasma* infection was considered asymptomatic. However, research over the past two decades indicates that individuals with latent toxoplasmosis exhibit a higher incidence of various diseases and generally score lower on health-related metrics compared to those without *Toxoplasma* [2]; for a review, see [3]. *Toxoplasma* has been also demonstrated to alter the behavior of its intermediate hosts. Examples of such behavioral alterations include a shift from a natural aversion to cat odors to attraction in laboratory-infected rodents [4,5], as well as prolonged reaction times in both infected rodents [6] and humans [7]. These changes have been hypothesized as being manipulations by the parasite to increase the likelihood of transmission to its definitive host, a cat, by predation. An alternative and likely simpler explanation is that these differences are merely side effects of the host’s deteriorated health. The side effects hypothesis was challenged by the results from path analysis in a recent study [8], showing that deteriorated health does not mediate the observed differences in psychological traits. It is crucial to note that this study specifically tested and refuted the mediating role of physical health solely for the psychological changes examined, not for cognitive impairments.

Compared to *Toxoplasma*, cytomegalovirus (CMV), a member of the herpesvirus family, infects even a larger proportion of the human population. The seroprevalence of CMV correlates with population age, being around 50% among university students and exceeding 80% in older demographics [9]. While latent postnatal CMV infection, akin to toxoplasmosis, was traditionally viewed as asymptomatic, emerging studies hint at its association with specific behavioral traits [10–16]. However, the behavioral impact of latent CMV infection has not received as much investigative attention as latent toxoplasmosis. Unlike toxoplasmosis, to date, there are no known behavioral experiments involving artificial CMV infection in laboratory animals, and longitudinal human studies are scarce [17,18]. Therefore, the causality relationship between infection and behavioral changes remains unresolved in the case of CMV.

Notably, CMV transmission does not occur through predation but through direct physical contact or body fluid exchange (e.g., during kissing). Based on evolutionary parasitological assumptions, this should lead to less specific and conspicuous expected behavioral modifications. This subtlety perhaps contributes to the topic’s lesser appeal for biologists interested in parasite manipulation. As of January 2024, the Web of Science database included 142 articles with ‘toxoplasm* AND behavio*’ in their titles, with 117 of these specifically addressing behavioral changes in latent toxoplasmosis. In contrast, the search query ‘(CMV OR cytomegalo*) AND behavio*’ yielded only 35 articles, and just four of these focused on postnatal CMV infection and its associated behavioral symptoms.

During the last 20 years of research into the effects of latent toxoplasmosis on human behavior and cognitive performance, it has been revealed that the Rh factor of the infected individual plays a significant role as a modifier of these impacts. Approximately 16% of Europeans are Rh-negative [19], a condition characterized by the absence of the immunodominant D epitope on the surface of erythrocytes. This absence is largely attributed to a near-complete deletion in the RHD gene [20]. The presence or absence of the RhD protein on erythrocytes is crucial in blood transfusion and maternal-fetal medicine, and has been a significant research focus for decades [21]. The RhD molecule is a component of the erythrocyte’s transmembrane protein-glycoprotein complex. It is predominantly recognized for its role in the transport of NH_3_, CO_2_, and their ions [22,23] and its role in the maintenance of plasma membrane integrity and the characteristic biconcave discoid shape of erythrocytes in humans [23,24]. However, the broader physiological implications of the Rh complex are not fully elucidated.

There is evidence that Rh-negative individuals may experience various health challenges, including cognitive and neurological issues [25–27]. Recent studies propose that Rh-negative individuals might face altered function of the ion channel containing two molecules of RhCE instead of one RhD and one RhCE molecule. This alteration could lead to localized hypoxia and heightened neuroinflammation in specific brain regions [27,28]. The resulting neurophysiological disturbances could potentially explain the observed association between Rh-negativity, toxoplasmosis, and certain neurological disorders [28].

Numerous studies have underscored that individuals with the Rh-positive phenotype, particularly heterozygotes with one D and one d allele of the RHD gene, exhibit increased resilience against negative influences like fatigue, smoking, and aging [29–34]. Intriguingly, this resilience also extends to *Toxoplasma* infection. Latent toxoplasmosis has been shown to have different effects on the behavior, performance, and health of individuals with Rh-negative and Rh-positive blood types [29]. For example, uninfected Rh-negative individuals usually display quicker reaction times compared to uninfected Rh-positive individuals. However, upon *Toxoplasma* infection, Rh-negative individuals often experience a substantial decline in reaction times. In contrast, Rh-positive heterozygotes tend to maintain or even improve their reaction times after infection [29,35]. The underlying mechanisms of this *Toxoplasma*-Rh interaction are still not understood, yet its existence has been corroborated by approximately ten studies over the last 15 years. This interaction also exhibits sex-specific variations; for more details, see [27,34]. No analogous effect has been noted in the context of human cytomegalovirus infection, highlighting the interesting interaction between *Toxoplasma* and the Rh blood group system. Nonetheless, it is pertinent to reiterate that significantly fewer studies have been conducted on the behavioral manifestations of CMV infection compared to the extensive research focusing on the behavioral effects of toxoplasmosis.

The general objective of our current study is to investigate the relationship of *Toxoplasma* or CMV infections with cognitive performance, respectively, in a cohort of 557 university students. Both pathogens are capable of surviving for decades in latent stages within the brain tissues of infected immunocompetent humans, where they can cause histopathological changes including localized inflammatory lesions [36,37]. Unlike *Toxoplasma*, which is transmitted trophically and may thus gain an advantage by reducing the vigilance and cognitive abilities of its intermediate host, CMV is not suspected of engaging in similar adaptive manipulation of host behavior and cognition. Therefore, by comparing cognitive performance deficits associated with *Toxoplasma* and CMV infections, we aim to discern whether these are solely the result of pathological changes in the brain, or also the outcome of deliberate manipulative activity by the pathogen to facilitate its transmission from infected to uninfected hosts. For this purpose, we utilized a comprehensive suite of tests assessing intelligence, knowledge, memory, simple reaction times, and information processing speed. Our analysis primarily aimed to assess the impact of *Toxoplasma* and CMV infections on human cognitive performance.

Additionally, in the exploratory part of the study, we investigated how sex and the Rh factor, recognized for their influence on toxoplasmosis’s behavioral effects, impact both infections. Specifically, for toxoplasmosis, we investigated how sex and Rh factor modulate specific aspects of cognitive performance. For CMV, the potential modulation effects of sex and Rh on cognitive outcomes had not been previously studied, thus our primary aim was to determine whether these factors influence the impact of infection on cognitive performance.

## 2. Methods

### 2.1. Subjects

We invited all undergraduate students enrolled in Evolutionary Biology and Practical Methodology of Science courses from 2010 to 2015 to participate voluntarily in a research project investigating the effects of parasites on human behavior, performance, and personality. The invitation was extended during lectures, and about 50% of the students consented, subsequently visiting our lab to provide 5ml of blood for serological analysis, collected by trained medical personnel. Owing to the extensive nature of the cognitive and reaction tests, which required approximately eleven hours per participant, our intention to keep the examination conditions identical in all rounds, and the limited capacity of our testing facility (maximum of 12 subjects simultaneously), the testing was conducted over an extended period. Intelligence, knowledge, and declarative memory assessments were carried out 2-24 months post blood collection. An additional 1-6 months later, participants returned for a third session to evaluate other memory types and reaction times. In total, data were collected from 557 students. During cognitive performance testing, neither the subjects nor the researchers were aware of the participants’ CMV and *Toxoplasma* infection statuses. Both male and female subjects were included in the study. Sex was identified based on participants’ responses to the demographic questionnaire query: ‘Your official sex at birth.’ The study adhered to the Declaration of Helsinki guidelines and received approval from the IRB of the Faculty of Science Charles University (protocol numbers 2009/06 and 2013/07). All participants were adults and provided written informed consent, as approved by the IRB.

### 2.2. Cognitive performance testing

#### 2.2.1. Intelligence

For intelligence assessment, we utilized Czech version of The Intelligence Structure Test I-S-T 2000 R [38,39]. This test is divided into two main modules: the basic module and the memory module. The basic module consists of three verbal, three numerical, and three abstract figural reasoning subtests. The memory module includes two memory tests and a knowledge test, which covers topics from geography and history, business, arts and culture, mathematics, science, and daily life. Employing both modules allowed us to comprehensively evaluate verbal, numerical, and figural intelligence, as well as verbal and figural memory, and verbal, numerical, and figural knowledge. These source variables were then used to calculate fluid intelligence, crystallized intelligence, general intelligence, and general knowledge (for further details, see the Memory subsection). Intelligence, memory, and knowledge scores (shown in Table 1) were derived from raw variables (i.e., the number of correct answers) using normative tables established for the entire Czech population. These tables do not account for age and sex; however, we controlled for these variables in our subsequent statistical analyses (refer to the Data Analysis section for details).

**Table 1.**
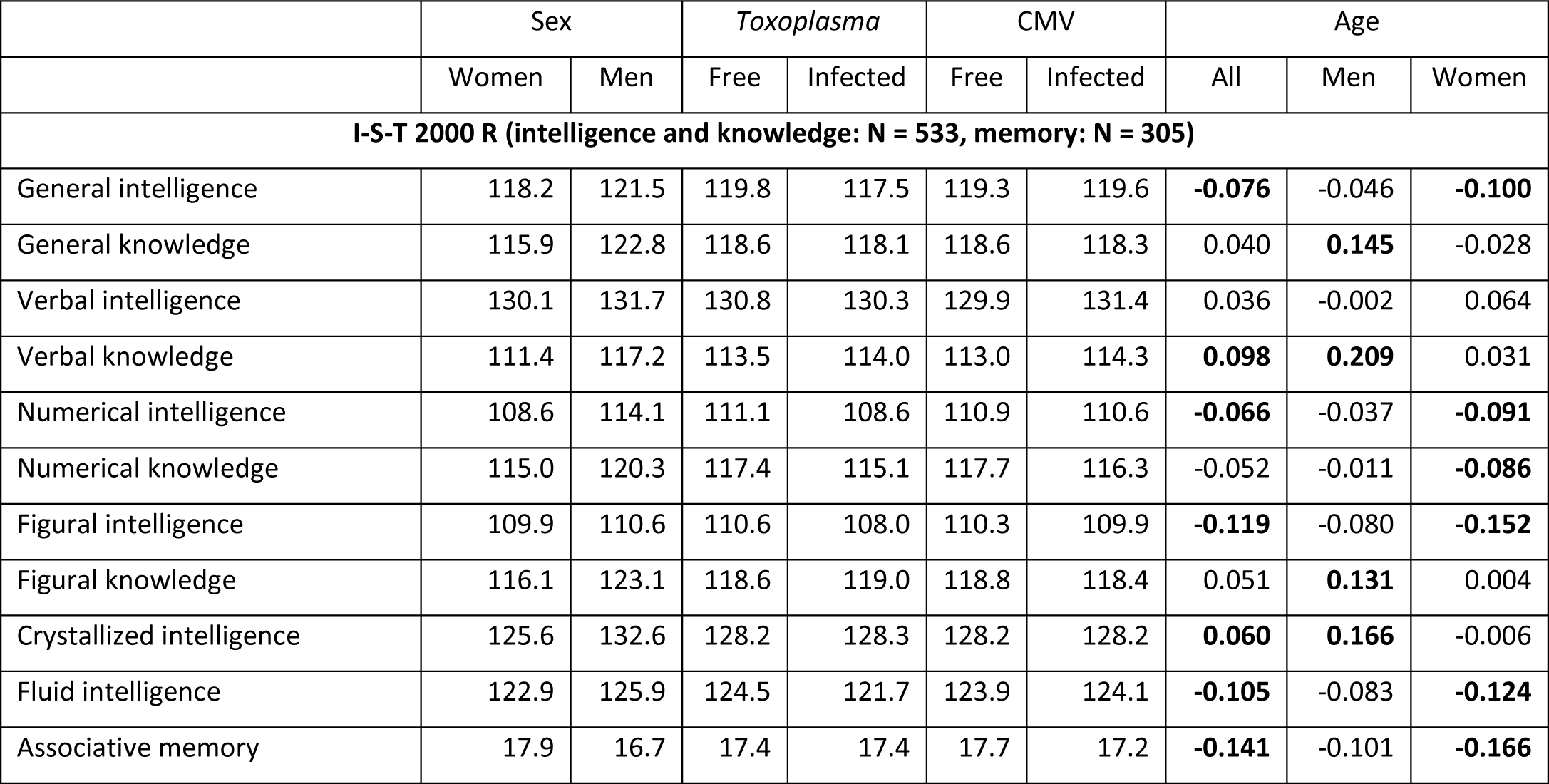
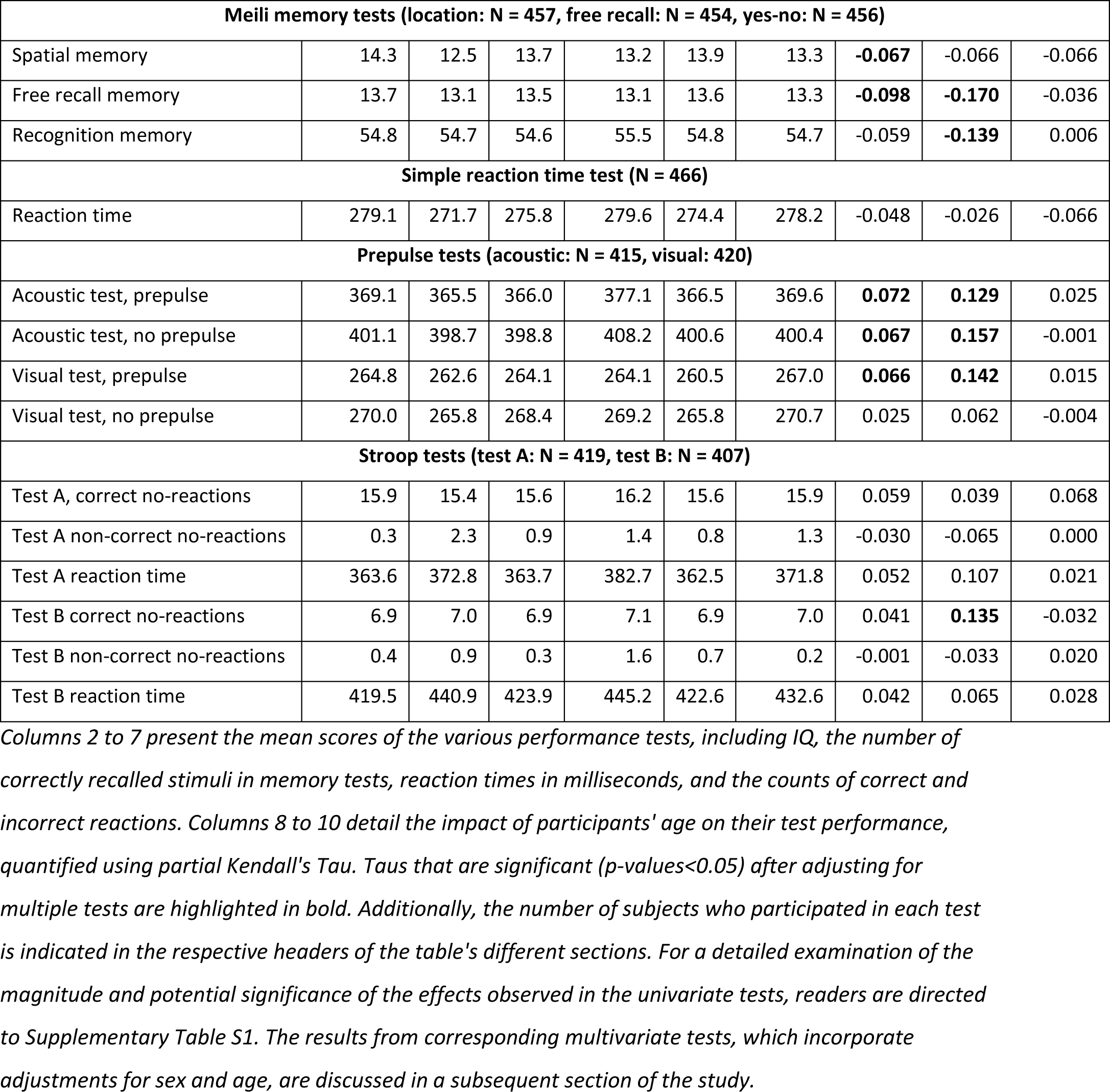
Average cognitive performance metrics by sex, *Toxoplasma* and CMV infection status, and age correlations.

The I-S-T 2000 R test was administered on computers to groups of 7–12 individuals simultaneously in the same room, with all sessions starting at 9:15 am. The total duration of the test was approximately 250 minutes, including a 15-minute break before the second module (comprising memory and knowledge tests). In conjunction with the I-S-T 2000 R session, participants also engaged in two physical performance tests, provided saliva samples for hormone concentration analysis, and underwent ECG recording. These additional data were utilized in an unrelated study [34].

#### 2.2.2. Memory

For the assessment of associative memory performance, we employed the I-S-T 2000 R memory module. The first part, the verbal memory test, involved a 60-second learning phase where participants were presented with 13 words across five categories, such as food, sports, and professions, each starting with a unique letter. Subsequently, they were asked to identify the category of a word beginning with a specified letter, such as ‘B’ (10 questions, maximum raw score of 10). The second part, the figural memory test, required participants to learn 13 pairs of abstract figures within 60 seconds. They were then shown 13 figures and given 3 minutes to match each figure with its pair from five options (13 questions, maximum raw score of 13). Associative memory was determined by summing correct answers from both tests, leading to a maximum raw score of 23. Due to technical issues, we were not able to administer the second part of the I-S-T 2000 R, which contains memory tests, to approximately one-third of the participants.

In the third testing session, we utilized the three-part Meili memory test [40] to evaluate other memory types, namely spatial memory, free recall memory, and recognition memory. Participants initially had 60 seconds to memorize 30 objects in a 6 × 5 grid of simple black and white drawings, such as a car, hat, candle, and flower [41]. Following a brief, intentional interruption of approximately one minute for the collection of their name, sex, and age, participants proceeded to complete a series of three subtests. In the first part, the spatial memory subtest, participants were presented with 30 drawings of the objects in succession, with each object displayed on a screen for a duration of 4 seconds. During these 4 seconds, participants were required to use a computer mouse to indicate the object’s original position on a blank 6 × 5 grid that was also displayed on the screen. There were no pauses between the presentations. The second part, lasting three minutes, involved the free recall memory subtest, where participants were prompted to write down as many shown objects as they could remember. The final part, the recognition memory subtest, presented each participant with 60 object names (30 original and 30 new), each for 4 seconds. They had to decide whether the picture of each object had been previously shown on the grid.

#### 2.2.3. Reaction times

In the simple reaction time test, participants were required to press the left button of a computer mouse in response to 40 simple visual go-stimuli. These stimuli appeared at irregular intervals of 2–8 seconds in the center of the computer screen. We excluded outlier values from the analysis, specifically reaction times longer than 700 milliseconds or shorter than 150 milliseconds [7]. However, it was noted that the results of all analyses were nearly identical even if this exclusion step was omitted.

The prepulse tests were designed similarly to the simple reaction time test, with the addition of random, low-intensity pulses occurring 20 milliseconds before half of the go-stimuli, accompanied by either acoustic or visual white noise. In the acoustic prepulse test, the stimuli, prepulses, and white noise were all acoustic. Conversely, in the visual prepulse test, these elements were visual. The visual white noise was represented by squares of varying shades of gray that randomly appeared and expanded on the screen. The visual prepulse test also included both prepulses in the screen’s central area and occasional ‘distractors’ – distinctive but brief signals – at various screen locations. In this study, we only used data on reaction times to stimuli with and without prepulses. The investigation of pathogens’ effect on prepulse inhibition/stimulation was addressed in a separate study [42].

#### 2.2.4. Information processing speed

To evaluate information processing speed and action control, we utilized two variants of the Stroop test. In both tests, participants were instructed to react swiftly to go-stimuli and to refrain from reacting to stop-stimuli. The stimuli were the words “black,” “green,” “red,” and “yellow,” displayed in various combinations of font color and word content on the computer screen. In the A variant of the Stroop test, participants were required to respond quickly to stimuli written in black, green, and red, but not to those in yellow. This version aimed to assess basic reaction time and inhibitory control. Conversely, the B variant was designed to test cognitive flexibility and the ability to adapt to changing rules: participants were to react to the same words in any font color except when the word was “yellow,” challenging their capacity to shift cognitive sets in response to altered task demands. Each variant included 100 stimuli. For all participants, we calculated the number of correct nonreactions (indicating action control), the number of incorrect nonreactions, and the mean reaction time of correct responses.

### 2.3. Immunological tests for *Toxoplasma*, CMV, and the Rh phenotype

*Toxoplasma* infection status was assessed at the National Reference Laboratory for Toxoplasmosis, National Institute of Public Health in Prague. We employed the complement fixation test and ELISA IgG test (both by TestLine Clinical Diagnostics) for this purpose. Subjects were considered non-infected with *T. gondii* if results from both tests were negative (CFT<8, ELISA < 175 OU), while those with positive results in both tests were considered *Toxoplasma* positive. Participants with one positive and one negative result were excluded from further analysis.

CMV status (infected vs. non-infected) was determined at the National Reference Laboratory for Herpes Viruses, National Institute of Public Health in Prague using the sandwich ELISA method with an inactivated CMV AD 169 strain antigen (ETI CYTOK G PLUS, DiaSorin, Saluggia, Italy), allowing quantitative detection of CMV IgG from 0.4 IU/ml to 10 IU/ml. Subjects with ambiguous serological test results (IgG concentration between 0.36–0.44 IU/ml) were also excluded.

The Rh phenotype, specifically the presence of the RhD antigen on erythrocyte membranes, was ascertained using a commercial agglutination test with human monoclonal anti-D reagents (Seraclone1, ImmucorGamma Inc.) during the blood collection session.

### 2.4. Data analyses

MANCOVA test was performed with software Statistica v.14. For the log-linear analysis of the 2 × 2 × 2 contingency tables (e.g., sex × infection × Rh table), we utilized an online calculator available at http://vassarstats.net/abc.html. All other statistical analyses, including path analysis, were conducted using R version 3.3.1 software [43], employing packages corpcor, pcaPP, tcltk2, ppcor, psych, lavaan, and semplot. Partial Kendall correlation tests were computed in R using the Explorer 0.1 program [44].

For an overall MANCOVA test, we employed Type VI sum of squares (unique SS) to determine the individual contribution of each independent variable, controlling for the effects of other variables in the model. This approach enabled us to isolate and assess the specific impact of each factor independently. Several variables related to cognitive performance displayed bimodal or multimodal distributions and varied between men and women, as well as between Rh-negative and Rh-positive individuals. They also showed strong correlations with participant age. Consequently, we employed nonparametric partial Kendall tests, controlling for age and sex, to investigate the effects of *Toxoplasma* and CMV infections, and the concentrations of specific antibodies in seropositive subjects (as a proxy for time since infection) on intelligence, knowledge, memory, reaction times, and action control. Previous studies have documented sex-dependent effects of *Toxoplasma* infection on various behavioral variables, many of which are also influenced by subjects’ Rh phenotype. For an extensive discussion on this topic, see [45]. Accordingly, our analyses were performed for the entire sample, and separately for men and women, and for Rh-negative and Rh-positive individuals. We controlled for the effect of multiple tests using the Benjamini-Hochberg procedure (FDR = 0.10) [46]. The complete dataset is available at Figshare: https://doi.org/10.6084/m9.figshare.21865677.

#### Technical Notes

Throughout the manuscript, we use the terms “toxoplasmosis” and “CMV infection” as shorthand for testing positive for the presence of anti-*Toxoplasma* or anti-CMV IgG antibodies in standard serological tests.

We consistently use the term “effect” in its statistical sense, referring to the difference between the true population parameter and the null hypothesis value. Only in the Discussion section do we distinguish between cause and effect, using “effect” in its non-technical sense.

Given the exploratory nature of the main part of this study, we report both the results corrected and non-corrected for multiple tests. For the same reason, in the exploratory section of the study, we discuss not only the statistically significant effects but also trends that did not reach formal significance.

## 3. Results

### 3.1. Descriptive statistics of the population under study

Our final dataset included 352 women (mean age 22.8, SD 3.6) and 205 men (mean age 24.2, SD 6.3). Although the age difference between women and men was small (Cohen’s d = 0.30), it was statistically significant (p = 0.004). In terms of infection prevalence, 51.8% of women and 51.5% of men were CMV positive, while toxoplasmosis prevalence was 17.4% in women and 17.6% in men. Fisher’s exact test revealed that the associations between sex and toxoplasmosis, sex and CMV, and toxoplasmosis and CMV were not statistically significant (p-values: 0.99, 0.93, and 0.57, respectively). In all groups, *Toxoplasma*- and CMV-infected subjects were generally older than their noninfected counterparts. However, this age difference was significant only for CMV-infected men (23.1 vs. 25.1, Cohen’s d = 0.32, p = 0.028).

The proportion of Rh-negative individuals was 19.3% in women and 19.1% in men, indicating no significant sex difference. In Rh-positive subjects, CMV infection rates were 51.7% for women and 55.4% for men, whereas in Rh-negative individuals, the rates were 52.3% for women and 33.3% for men. Following this observation, a log-linear analysis of the 2 × 2 × 2 contingency table (sex × infection × Rh) indicated that the interaction between Rh factor, CMV infection, and sex was not statistically significant (G^2^ = 5.8, degrees of freedom = 2, p = 0.055). Given the number of tests conducted, the observed difference in CMV prevalence among Rh-negative men and women is likely due to chance. No similar trend was observed with *Toxoplasma* (G^2^ = 3.16, degrees of freedom = 2, p = 0.206).

Descriptive statistics for variables related to cognitive performance, as well as the correlations of these variables with subjects’ age, are detailed in Table 1. This table also includes the number of students who participated in each specific performance test.

### 3.2. Associations of toxoplasmosis, CMV infection, Rh phenotype, sex, and their interactions with cognitive performance

Before proceeding with more detailed analyses, it was necessary to determine whether *Toxoplasma* and CMV infections have a discernible impact on cognitive performance and if this effect is modulated by the Rh phenotype and sex. Therefore, we began our analysis with an overall MANCOVA test. The dependent variables were the results of all performance tests (see Table 1), with toxoplasmosis, CMV infection, Rh phenotype, and sex as binary independent variables, and age as a continuous covariate. The results are presented in Table 2. For toxoplasmosis, only the main effect was significant (p = 0.040, partial eta² = 0.175). For CMV infection, the main effect was of similar strength but non-significant (p = 0.071, partial eta² = 0.164) and significant effects were observed for the double interactions of sex-CMV (p = 0.002, partial eta² = 0.223) and CMV-Rh (p = 0.022, partial eta² = 0.185), as well as the triple interaction of sex-CMV-Rh (p = 0.002, partial eta² = 0.223). The direction of these effects for each variable will be elucidated in the exploratory section of our study, providing a detailed understanding of how these factors individually and collectively impact cognitive performance. Although interactions of toxoplasmosis with Rh and sex were not significant this time, based on the results of previous studies, we decided to analyze the effects of both pathogens in the same manner in the subsequent exploratory part of the study, which focused on the influence of these factors on individual facets of cognitive performance. Due to the asymmetric distribution of some test outcome variables, significant differences in the number of individuals in various subpopulations, and relatively low counts in some subpopulations, we employed partial Kendall correlation tests in post-hoc analyses, which are not sensitive to these issues.

**Table 2.** Associations of various biological factors with cognitive performance - multivariate MANCOVA test.

|  | **F<sub>25,190</sub>** | **p** | **Partial eta<sup>2</sup>** |
| --- | --- | --- | --- |
| Intercept | 933.7954 | **0.000** | 0.992 |
| sex | 2.0472 | **0.004** | 0.212 |
| tox | 1.6092 | **0.040** | 0.175 |
| cmv | 1.4916 | 0.071 | 0.164 |
| Rh | 0.8422 | 0.684 | 0.100 |
| sex*tox | 0.8322 | 0.697 | 0.099 |
| sex*cmv | 2.1844 | **0.002** | 0.223 |
| tox*cmv | 1.1808 | 0.261 | 0.134 |
| sex*Rh | 0.8172 | 0.717 | 0.097 |
| tox*Rh | 1.2188 | 0.227 | 0.138 |
| cmv*Rh | 1.7244 | **0.022** | 0.185 |
| sex*tox*cmv | 0.9333 | 0.559 | 0.109 |
| sex*tox*Rh | 0.8077 | 0.729 | 0.096 |
| sex*cmv*Rh | 1.6449 | **0.033** | 0.178 |
| tox*cmv*Rh | 0.9956 | 0.475 | 0.116 |
| sex*tox*cmv*Rh | 1.0808 | 0.368 | 0.125 |
| age | 2.9912 | **0.000** | 0.282 |

*The table presents the outcomes of the MANCOVA test (using Type VI sum of squares) that included the results of all performance tests as dependent variables. Age was incorporated as a covariate, while toxoplasmosis, CMV infection, Rh phenotype, and sex were used as independent variables. Statistically significant effects are highlighted in bold. For clarity, significance levels (p-values) lower than 0.0005 are reported as 0.000 in the table.*

### 3.3. The association of *Toxoplasma* and CMV infections with different facets of cognitive performance

We examined the association between cognitive performance test outcomes and *Toxoplasma* and CMV infections using partial Kendall correlation tests, adjusting for age and sex variables. Table 3 presents the effect sizes (partial Kendall Tau) and their significance, both before and after adjustments for multiple tests. Our results reveal that toxoplasmosis significantly impacts various facets of intelligence in men, specifically numerical, figural, fluid, and general intelligence, as well as several psychomotor performance metrics. In contrast, CMV infection did not demonstrate a notable effect on intelligence measures. However, it notably influenced memory, as determined by three distinct memory tests, with significant findings particularly among women. Additionally, CMV infection was found to affect reaction times and action control in a way similar to *Toxoplasma* infection, though these effects were more pronounced in women compared to men.

**Table 3.** Association of *Toxoplasma* and CMV infections with performance in cognitive tests.

|  | All *Toxoplasma* |  | Women *Toxoplasma* |  | Men *Toxoplasma* |  | All CMV |  | Women CMV |  | Men CMV |  |
| --- | --- | --- | --- | --- | --- | --- | --- | --- | --- | --- | --- | --- |
|  | Infection | IgG | Infection | IgG | Infection | IgG | Infection | IgG | Infection | IgG | Infection | IgG |
| **I-S-T 2000 R** |  |  |  |  |  |  |  |  |  |  |  |  |
| General Intelligence | **-0.071** | -0.031 | -0.015 | -0.021 | **-0.161** | -0.075 | 0.011 | 0.062 | 0.023 | 0.055 | -0.006 | 0.059 |
| General knowledge | -0.029 | -0.058 | 0.019 | -0.047 | **-0.124** | -0.069 | -0.011 | -0.014 | -0.041 | 0.001 | -0.016 | -0.082 |
| Verbal intelligence | -0.005 | -0.003 | 0.004 | 0.036 | -0.014 | -0.080 | 0.053 | 0.020 | 0.047 | 0.013 | 0.055 | 0.027 |
| Verbal knowledge | 0.007 | -0.037 | 0.029 | -0.059 | -0.037 | -0.001 | 0.046 | -0.018 | 0.017 | -0.034 | 0.077 | -0.001 |
| Numerical intelligence | **-0.059** | -0.001 | 0.012 | -0.017 | **-0.160** | 0.002 | 0.004 | 0.054 | 0.013 | 0.073 | -0.004 | 0.021 |
| Numerical knowledge | **-0.062** | -0.094 | -0.018 | -0.112 | **-0.133** | -0.089 | -0.042 | -0.001 | -0.071 | 0.023 | -0.022 | -0.044 |
| Figural intelligence | **-0.058** | -0.021 | -0.022 | -0.005 | **-0.121** | -0.048 | -0.004 | 0.044 | 0.005 | 0.016 | -0.027 | 0.102 |
| Figural knowledge | 0.002 | -0.007 | 0.042 | 0.089 | -0.082 | -0.166 | -0.014 | -0.032 | -0.024 | -0.028 | -0.007 | -0.056 |
| Crystallized intelligence | -0.010 | -0.054 | 0.020 | -0.056 | -0.079 | -0.069 | -0.002 | -0.040 | -0.034 | -0.034 | 0.013 | -0.090 |
| Fluid intelligence | **-0.063** | -0.011 | -0.001 | 0.003 | **-0.165** | -0.055 | 0.010 | 0.062 | 0.016 | 0.072 | 0.007 | 0.042 |
| Associative memory | -0.038 | 0.038 | -0.040 | 0.183 | -0.050 | -0.153 | -0.075 | 0.007 | **-0.099** | 0.021 | -0.049 | -0.056 |
| **Meili memory tests** |  |  |  |  |  |  |  |  |  |  |  |  |
| Spatial memory | -0.041 | -0.036 | -0.061 | 0.036 | -0.017 | -0.141 | -0.060 | 0.037 | **-0.149** | -0.011 | 0.086 | 0.108 |
| Free recall | -0.043 | -0.086 | -0.072 | 0.043 | 0.003 | **-0.307** | -0.061 | 0.015 | **-0.111** | -0.014 | 0.029 | 0.051 |
| Recognition memory | 0.061 | -0.003 | 0.056 | 0.074 | 0.075 | -0.154 | -0.025 | 0.045 | -0.070 | 0.000 | 0.055 | 0.083 |
| **Simple reaction time test** |  |  |  |  |  |  |  |  |  |  |  |  |
| Reaction time | 0.027 | -0.067 | -0.012 | 0.100 | 0.086 | -0.003 | **0.071** | -0.033 | **0.100** | 0.037 | 0.025 | **-0.157** |
| **Prepulse tests** |  |  |  |  |  |  |  |  |  |  |  |  |
| Acoustic test, prepulse | 0.064 | **-0.174** | 0.064 | **-0.206** | 0.057 | -0.182 | 0.040 | 0.035 | 0.042 | 0.107 | 0.028 | -0.041 |
| Acoustic test, no prepulse | 0.048 | -0.108 | 0.023 | **-0.201** | 0.085 | -0.016 | 0.036 | 0.028 | 0.039 | 0.081 | 0.028 | -0.030 |
| Visual test, prepulse | -0.005 | -0.068 | -0.012 | -0.139 | 0.006 | 0.062 | **0.090** | 0.039 | **0.083** | 0.070 | 0.101 | 0.020 |
| Visual test, no prepulse | 0.006 | -0.026 | -0.010 | -0.054 | 0.025 | 0.013 | **0.068** | 0.014 | 0.061 | 0.045 | 0.081 | -0.005 |
| **Stroop tests** |  |  |  |  |  |  |  |  |  |  |  |  |
| Test A, correct no-reactions | 0.052 | -0.001 | -0.046 | 0.028 | <u>**0.205**</u> | -0.085 | 0.021 | 0.027 | <u>**0.154**</u> | 0.030 | <u>**-0.177**</u> | 0.033 |
| Test A incorrect no-reactions | 0.059 | -0.079 | 0.058 | -0.126 | 0.060 | -0.038 | 0.052 | 0.089 | 0.049 | 0.044 | 0.062 | 0.141 |
| Test A reaction time | <u>**0.129**</u> | -0.124 | **0.091** | -0.095 | <u>**0.207**</u> | -0.162 | <u>**0.117**</u> | 0.013 | <u>**0.165**</u> | 0.001 | 0.027 | 0.034 |
| Test B correct no-reactions | 0.063 | -0.088 | 0.028 | 0.028 | <u>**0.124**</u> | -0.245 | 0.001 | -0.030 | 0.005 | 0.029 | -0.019 | -0.112 |
| Test B incorrect no-reactions | 0.028 | 0.023 | -0.010 | -0.108 | 0.084 | 0.202 | 0.019 | **-0.099** | 0.053 | -0.103 | -0.030 | -0.077 |
| Test B reaction time | <u>**0.116**</u> | **-0.178** | 0.082 | -0.153 | <u>**0.188**</u> | -0.243 | <u>**0.098**</u> | -0.022 | <u>**0.141**</u> | 0.038 | 0.029 | -0.101 |

*This table delineates the magnitude and direction (partial Kendall's Tau) of the effects of infection status (binary variable: 0 for infection-free, 1 for infected) and specific antibody concentrations (quantified via ELISA; analysis of antibody concentrations was restricted to infected subjects) on participants' performance in various cognitive tests. The statistical tests were controlled for age and sex variables. Statistically significant effects are highlighted in bold. Additionally, those effects that retain their significance after correction for multiple tests are marked with an underline.*

### 3.4. Correlations between the concentrations of specific anti-*Toxoplasma* and anti-CMV antibodies and cognitive performance

Table 3 also shows the effects of anti-*Toxoplasma* IgG and anti-CMV IgG antibody concentrations on the cognitive performance of infected subjects. Within the context of toxoplasmosis, specific IgG antibody levels may act as a proxy for the time elapsed since infection [47], a hypothesis we also tested for CMV. In *Toxoplasma*-infected men, high concentrations of anti-*Toxoplasma* antibodies, characteristic of the disease’s earlier stages, were associated with reduced intelligence, impaired memory, and faster reaction times in most tests, suggesting that as the infection progresses beyond its acute phase, the negative effects on intelligence and memory tend to decrease, whereas the influence on psychomotor performance, evidenced by prolonged reaction times, tend to increase. However, these associations, though relatively strong, did not achieve statistical significance, likely due to the limited number of infected male subjects in our study. In women, a similar trend was observed, but the effects were generally milder. An exception was found in the acoustic prepulse test, where a stronger and significant negative correlation between antibody concentration and reaction time was noted in women compared to men. The most notable sex difference emerged in memory test performance: men exhibited improved performance with decreasing anti-*Toxoplasma* IgG antibodies, presumably as time since infection increased, whereas women showed decreased performance with lower antibody levels.

The associations between anti-CMV antibodies and cognitive performance were considerably weaker and did not display a consistent pattern for either sex. However, when considering the modifying impact of the Rh phenotype, a more distinct pattern began to emerge, as will be discussed in subsequent sections.

### 3.5. The mechanism of improved action control in *Toxoplasma*- or CMV-infected individuals

Previous studies on toxoplasmosis have indicated that *Toxoplasma*-infected subjects exhibit superior action control, defined as the ability to inhibit a response to a false stimulus, compared to noninfected individuals [48]. In line with this, our data revealed that men infected with *Toxoplasma* and women infected with CMV displayed significantly longer reaction times to go-stimuli, but committed fewer errors in responding to stop-stimuli, as shown in Table 2. To explore whether prolonged reaction times could be a proximate cause of the reduced error rate in infected subjects, we employed structural equation modeling, specifically path analysis (PA). The PA results, illustrated in Figure 1, indeed showed low (and negative) path coefficients linking infection status to the number of correct non-reactions, and high, significant positive path coefficients connecting reaction times to correct non-reactions.

**Figure 1.**
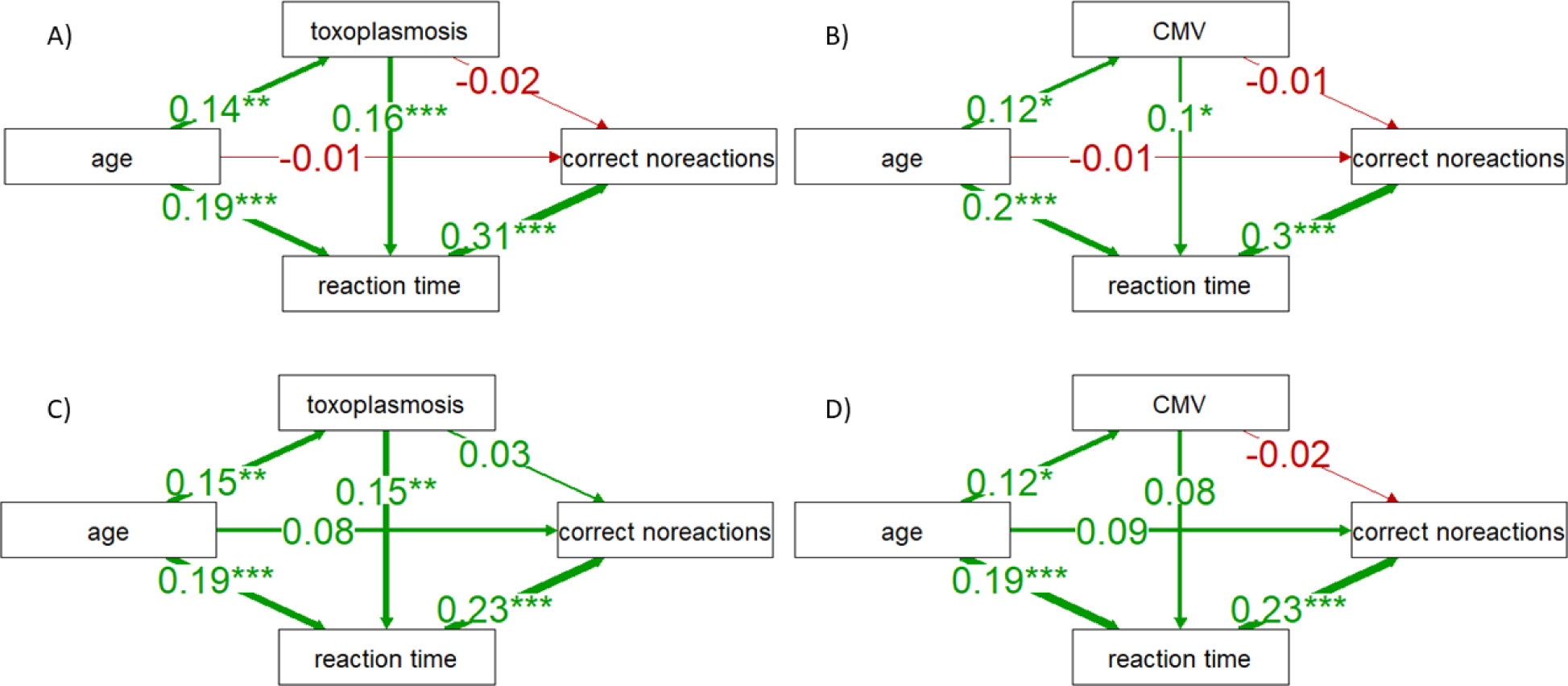
The results of path analyses showing direct and reaction time-mediated effects of *Toxoplasma* and CMV infections on subjects’ performance in the Stroop tests. Schemes A and B show the results (path coefficients) for Stroop test A; schemes C and D show the same for Stroop test B. One, two, and three asterisks indicate associations at the levels of 0.05, 0.01, and 0.001, respectively.

### 3.6. Modifying effects of the Rh phenotype

To further explore, we conducted partial Kendall correlation tests controlled for age separately for Rh-positive and Rh-negative men and women. The outcomes of these analyses for *Toxoplasma* and CMV infections are presented in Table 4. It is important to note that the number of Rh-negative subjects in our study, which mirrored the natural occurrence of Rh-negative individuals in the population, was four times lower than that of Rh-positive subjects. As a result, many of the effects in the Rh-negative subgroup did not achieve formal statistical significance. However, the effect sizes in Rh-negative individuals were generally, albeit not consistently, larger than those in Rh-positive individuals. Notably, the patterns of these effects varied depending on the type of infection (*Toxoplasma* or CMV), sex of the subjects, and across different cognitive performance tests.

**Table 4.**
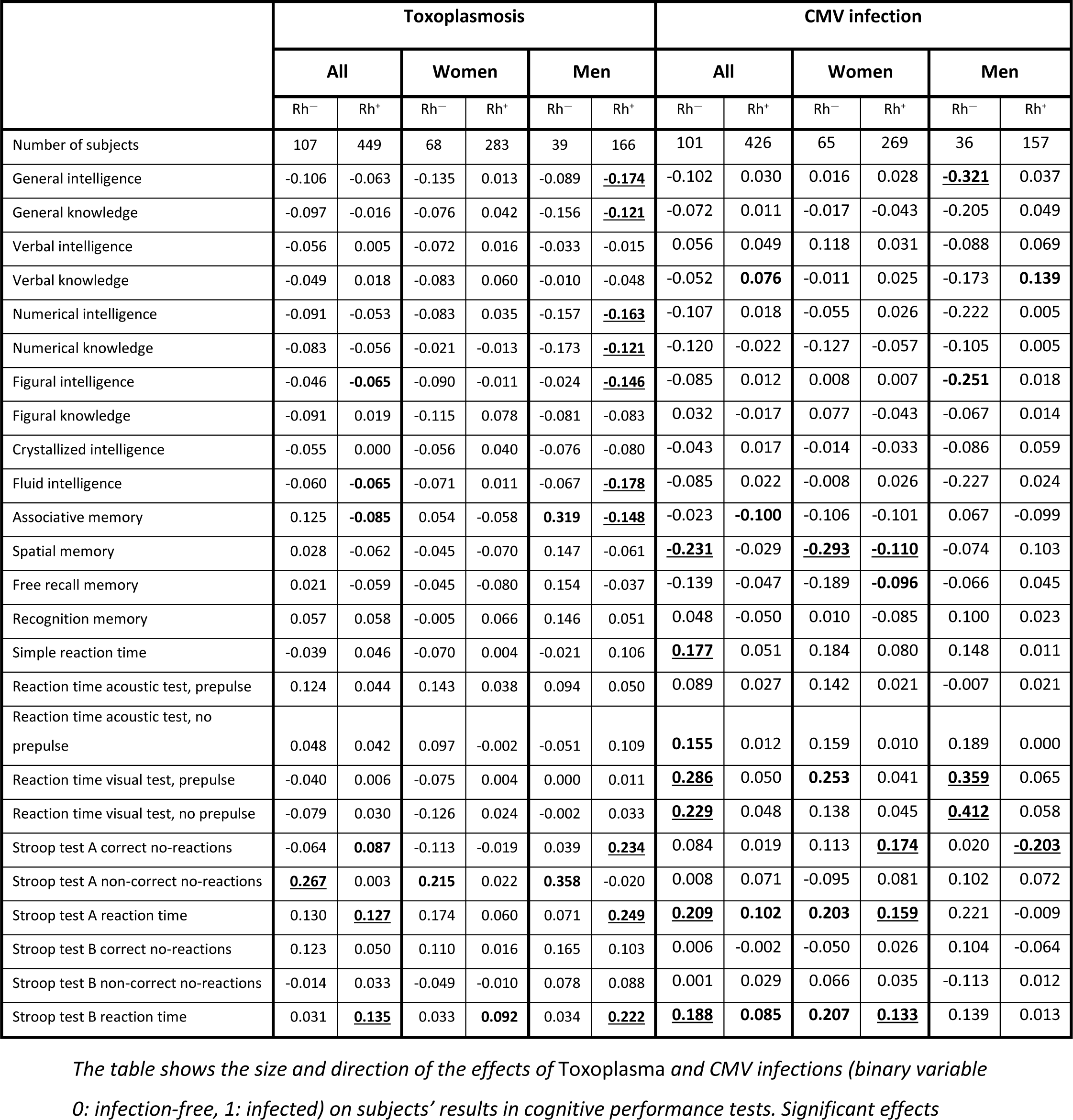
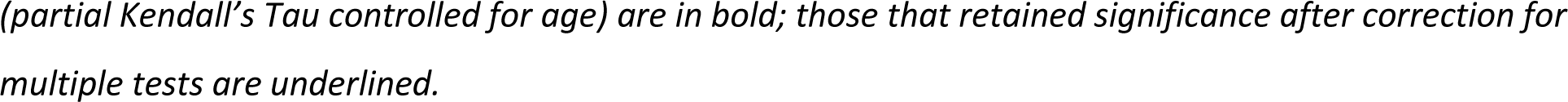
Association of *Toxoplasma* and CMV infections with performance in cognitive tests in Rh-positive (Rh+) and Rh-negative (Rh^—^) subjects.

## 4. Discussion

### 4.1. Confirmatory part of the study

The primary aim of our study was to compare the effects of two neurotrophic pathogens—*Toxoplasma*, a parasite known to manipulate its host’s behavior, and human CMV, a virus not associated with behavioral manipulation—on various cognitive functions. We aimed to evaluate their influence on intelligence, memory, reaction times, and information processing speed, as measured through a series of cognitive tests, with the assumption that *Toxoplasma* would have a more pronounced or evident effect. Contrary to our expectations that significant cognitive impairments would be associated solely with Toxoplasma, our findings revealed that both Toxoplasma and CMV were associated with predominantly adverse effects of similar magnitude on the cognitive performance of subjects. The strength of the associations of *Toxoplasma* and human CMV with various cognitive functions varied depending on the pathogen involved and was influenced by the sex and Rh factor status of the individuals. Specifically, the association with toxoplasmosis was more pronounced and widespread in men, while the association with CMV infection was stronger and more prevalent in women. In line with previous research [29–34], the associations were typically stronger in Rh-negative individuals and either weaker or non-existent in Rh-positive individuals.

The formation of lifelong dormant stages capable of reactivation in the brain during immunosuppression or following a head injury is a characteristic shared by both *Toxoplasma* and CMV [49–52]. However, modes of transmission of *Toxoplasma* and CMV differ. While *Toxoplasma* is transmitted through predation [53], a process that could potentially be facilitated by the impairment of a host’s cognitive abilities, CMV is transmitted through direct contact, such as saliva, urine, milk, and genital secretions [54] and is not presumed to manipulate host behavior to enhance its transmission. Given that we observed lower cognitive performance in individuals infected with *Toxoplasma*, as well as those with CMV, it seems plausible that the differences noted between infected and non-infected subjects are more likely a result of pathological processes occurring locally in the brain due to the presence of dormant stages of both pathogens [36,37], rather than a result of their manipulative activity aimed at facilitating transmission from infected to uninfected hosts. However, it is important to acknowledge that this suggestion does not constitute definitive proof but rather indicates a possible direction for further investigation.

### 4.2. Exploratory part of the study

#### 4.2.1. Intelligence and toxoplasmosis

Our study indicated that toxoplasmosis negatively influenced several aspects of intelligence, encompassing general, numerical, figural, and fluid intelligence. Notably, fluid intelligence, associated with problem-solving abilities, comprehension, and reasoning, was significantly affected. In contrast, its counterpart, crystallized intelligence—which relies on the recall of stored knowledge and past experiences—did not exhibit notable differences between *Toxoplasma*-infected and non-infected individuals. The adverse effects of toxoplasmosis on intelligence were primarily observed in men, while women exhibited either weaker or no such effects. Intriguingly, the Rh factor played only a minor role in the association between *Toxoplasma* infection and intelligence, unlike its impact observed in other performance tests or with CMV infection. The associations in Rh-negative and Rh-positive subjects largely differed in magnitude rather than direction. It’s important to mention that the effect sizes, indicated by Tau values, were typically larger in Rh-negative subjects; however, the sample size in this group was four times smaller, which inevitably limited the power of the tests and thus our chance to detect significant results.

Our study did not find a formally significant effect of anti-*Toxoplasma* IgG concentration on the intelligence of *Toxoplasma*-infected students, yet all observed trends were negative, indicating a negative association between the levels of anti-*Toxoplasma* IgG concentration and intelligence. This suggests that intelligence levels may be lowest shortly after infection, coinciding with the highest concentration of anti-*Toxoplasma* antibodies, and potentially improve over time post-infection. This pattern aligns with previous findings showing a negative correlation between anti-*Toxoplasma* antibody concentration and intelligence, which was significant in that study [55]. Such correlation suggests that the observed decrease in intelligence among *Toxoplasma*-positive men could be temporary, stemming from the acute phase of the infection, rather than being due to the cumulative effects of latent toxoplasmosis. However, an alternate explanation for the negative trends is that the antibody concentration may reflect not only the time elapsed since acquiring toxoplasmosis but also the intensity of the infection, which could correlate with the extent of brain damage.

The impact of toxoplasmosis on intelligence remains a complex subject. While certain studies have suggested a negative influence on performance in IQ tests, others have reported contrasting results [55–57]. This variation in findings could be attributed to the wide range of both direct and indirect effects that *Toxoplasma* infection has on human behavior and physiology. Beyond its impact on intelligence, the infection has been linked to psychological traits like increased competitiveness, possibly due to increased testosterone levels in men [35,58], and altered levels of cooperativeness—decreasing in men while increasing in women [59]. Both competitiveness and cooperativeness can significantly affect outcomes on IQ tests. For certain subpopulations and in specific aspects of intelligence, such as verbal intelligence, the positive influences of these traits might counterbalance the detrimental effects of the infection on cognitive performances, including those on IQ tests [45].

#### 4.2.2. Intelligence and CMV infection

The effects of CMV infection on intelligence and cognitive performance, particularly in young and middle-aged individuals, have not been extensively studied. An earlier study [60], which stemmed from the first author’s diploma thesis [61], reported negative impacts of CMV infection on the IQ of university students. However, this study was retracted due to an error in the permutation tests, specifically the mislabeling of infected and uninfected individuals’ codes [62].

The present study found many adverse effects of CMV infection on various facets of intelligence. These effects were particularly pronounced in Rh-negative men, however, due to the low representation of Rh-negative individuals in the sample (19%) and the underrepresentation of men among university students (36.7%), the size of this specific group was relatively small (24 CMV-free and 12 CMV-infected individuals). Consequently, most of these substantial effects remained formally nonsignificant in the Rh-negative subpopulation, with the exception of the negative impacts on general and figural intelligence. In this context, it’s worth noting that the partial Kendall correlation is an ‘exact test’, a term referring not only to its precision but also to a category of statistical analyses that are robust even with limited or unevenly distributed data.

In contrast to the findings related to *Toxoplasma*, the analysis did not reveal any consistent pattern in correlations between anti-CMV IgG antibody concentration and the intelligence of CMV-infected subjects. We observed an equal number of positive and negative trends. Such a pattern might be expected if the decline in intelligence post-infection was relatively rapid and concluded within a few years or even months following CMV infection. However, it is crucial to remember that, unlike with toxoplasmosis [47,57,63], no study to date has shown a correlation between anti-CMV IgG antibody concentration and time since infection. In fact, some evidence suggests that anti-CMV IgG levels may increase over time post-initial CMV infection, or at least with the age of the patients [64,65].

#### 4.2.3. Toxoplasmosis and memory

Research into the effects of toxoplasmosis on memory functions in young and healthy individuals has been relatively scarce compared to its effects on intelligence. A recent meta-analysis [66] reviewed six articles addressing the impact of toxoplasmosis on working memory, revealing an aggregate effect size (Standardized Mean Difference, SMD) of 0.16 with a p-value of 0.002. Additionally, it identified five articles that studied short-term word memory, showing an aggregate effect of 0.18 with a p-value of less than 0.001. However, except for one study [67], these analyses predominantly focused on senior or middle-aged populations.

In our current study, we assessed the memory of *Toxoplasma*-infected and *Toxoplasma*-free university students using four different memory tests. Our findings indicated mostly adverse, yet nonsignificant, effects of toxoplasmosis on the students’ memory performance. The lack of a significant overall effect on memory might be attributed to toxoplasmosis having opposite impacts on Rh-positive and Rh-negative subjects. Notably, this effect was more pronounced in male students: Rh-positive infected males tended to perform worse, while Rh-negative infected males performed better than their *Toxoplasma*-free male counterparts in three out of the four memory tests. Again, due to the lower prevalence of Rh-negative subjects in our study sample, these observed effects were significant only in the I-S-T 2000 R memory test and not in any variant of the Meili tests. Similar patterns were observed in female students, although the effects were weaker and did not reach statistical significance.

#### 4.2.4. CMV and memory

Research on the impact of CMV infection on memory functions has predominantly focused on patients with dementia or seniors [11,68-70]; however, some studies have deviated from this trend [13,71]. For instance, a longitudinal study over five years involving 1,022 older individuals from the Monongahela– Youghiogheny Healthy Aging Team assessed cognitive decline annually using various cognitive tests, including memory tests. This study revealed that CMV-infected individuals experienced a significantly greater cognitive decline over time compared to their non-infected counterparts [18]. On the contrary, Gale et al. [72] reported no association between CMV and memory in a community-based sample of adults aged 40 to 70 years.

In our current study, we found that CMV-infected female students, unlike their male counterparts, performed significantly worse than non-infected peers in three of the four memory tests. Further analysis, stratified by Rh factor, showed that this effect was much more pronounced in Rh-negative women.

#### 4.2.5. Toxoplasmosis and reaction times

The first study reporting the effects of toxoplasmosis on reaction times was published in 2001 [7]. It showed that *Toxoplasma*-infected blood donors had longer reaction times than *Toxoplasma*-free blood donors and that in *Toxoplasma*-infected individuals, reaction times correlate negatively with concentrations of anti-*Toxoplasma* IgG antibodies (which decrease with time passed since infection). This suggests that reaction times worsen as more time elapses since the acute phase of toxoplasmosis. This could indicate that rather than being aftereffects of acute toxoplasmosis, we are seeing a cumulative effect of latent toxoplasmosis.

Later studies demonstrated that the effect of toxoplasmosis on the reaction times of blood donors and students depends on the subjects’ Rh factor status [29,35]. Rh-negative *Toxoplasma*-free subjects (Toxo-Rh-) had the shortest reaction times and Rh-negative *Toxoplasma*-infected subjects (Toxo+Rh-) had the longest reaction times from all four groups (Toxo+Rh+, Toxo+Rh-, Toxo-Rh+, Toxo-Rh-) of subjects. Moreover, the results showed that the effect of toxoplasmosis depended on the Rh genotype rather than Rh phenotype – the protective effect of Rh-positivity against the effects of toxoplasmosis was much stronger in Rh-positive heterozygotes than in Rh-positive homozygotes [29]. These findings, among others, have prompted the theory that Rh polymorphism may be maintained in human populations through selective advantages conferred upon heterozygotes in environments where *Toxoplasma* infection is common, which, until recently, was nearly ubiquitous [26,27,29,35].

In our current study, we found no substantial evidence to suggest any effect of toxoplasmosis on reaction times as measured by the simple reaction time test. However, the prepulse tests revealed predominantly negative effects (or trends) in relation to the concentration of anti-*Toxoplasma* IgG antibodies on reaction times. This pattern implies that reactions tend to become slower as more time passes since the infection. Such findings suggest that the cumulative effects of latent toxoplasmosis, rather than diminishing effects of past acute toxoplasmosis, might be contributing to the observed changes in reaction times.

Moreover, the Stroop tests indicated a significant detrimental effect of toxoplasmosis on reaction times, with more pronounced effects observed in men and variations noted between Rh-positive and Rh-negative individuals. However, these effects were not consistent across the two versions of the Stroop test, introducing an element of uncertainty in the interpretation of these results. Similar to the prepulse tests, the Stroop tests also exhibited negative effects (or strong, though nonsignificant trends) of the concentrations of anti-*Toxoplasma* IgG antibodies on reaction times.

#### 4.2.6. Toxoplasmosis and action control

An essential outcome from the Stroop tests in our study was the measurement of the number of ‘correct nonreactions’. Notably, *Toxoplasma*-infected men, but not women, exhibited better scores in this metric compared to *Toxoplasma*-free controls. Further analysis, stratified by Rh factor, indicated that Rh-negative individuals, particularly men, were primarily responsible for this effect. A higher number of correct nonreactions is typically interpreted as indicative of better action control, meaning the subject’s ability to halt an already initiated reaction to a false stimulus.

Previous studies have reported and discussed the enhanced performance of *Toxoplasma*-infected subjects in tasks requiring strong action control [48,73]. However, the results of our path analyses suggest that this improved performance in the test might be a secondary effect of their longer (worse) reaction times. In essence, the infected subjects do not necessarily exhibit better action control; rather, they have more time to adjust their reaction due to slower responses to stimuli.

#### 4.2.7. CMV and reaction times

CMV-infected subjects, particularly women, exhibited worse performance on nearly all reaction time tests, with the exception of the acoustic prepulse tests. This effect was more pronounced in Rh-negative individuals compared to Rh-positive ones. Specifically, in the first Stroop test (version A), CMV-infected women demonstrated a higher number of correct non-reactions. However, among men, those who were Rh-positive and CMV-infected scored lower in action control compared to their Rh-positive, CMV-free counterparts.

From a broader perspective, the observed impact of CMV infection on reaction times and cognitive performance introduces significant skepticism towards the hypothesis that exclusively associates the existence of Rh polymorphism in Europe with historical fluctuations in toxoplasmosis prevalence [29,35]. Our findings, by demonstrating comparable Rh-dependent interactions in the context of CMV infection, suggest that attributing Rh polymorphism solely to shifts in toxoplasmosis prevalence may be an oversimplification or potentially incorrect.

### 4.3. Strength and limitations

This study exhibits several significant strengths as well as certain limitations. A primary strength, particularly in comparison with similar studies as underscored in a recent meta-analysis [66], is the considerable number of participants involved. This larger sample size significantly enhanced the statistical power of our analyses. However, the stratified analyses encountered challenges, especially due to the small sizes of certain subgroups, such as Rh-negative *Toxoplasma*-infected men, as outlined in Table 3. As a result, the findings from some Rh-stratified analyses should be approached with caution. Additionally, the limited representation of specific participant classes in our sample restricted the ability to include more factors in our analytical models. For example, the current sample size did not permit a thorough investigation into the effects of CMV-*Toxoplasma* and CMV-*Toxoplasma*-Rh interactions on cognitive performance. This underscores the need for future studies with larger and more diverse cohorts to delve deeper into these intricate interactions and their impact on cognition.

Previous research on the effects of latent toxoplasmosis on intelligence often relied on relatively simple IQ tests, focusing primarily on facets like verbal or figural intelligence [55,56,74]. The key advantage of our current study is the use of the comprehensive I-S-T 2000 R questionnaire, a four-hour-long psychometric tool that measures or calculates a wide range of intelligence facets, as well as knowledge and memory.

A particular consideration in our study was the time gap between performance testing and serological testing for pathogens in some students. While this is less of a concern for *Toxoplasma*, due to the relatively low incidence of infection in this age group [75,76], it poses a potential issue for CMV. The incidence of CMV infections is relatively high in students, and it is conceivable that an unknown number of them might have acquired a CMV infection during the period between the serological and performance tests. The inclusion of such CMV-positive individuals with initially negative serological test results could have attenuated the observed differences between CMV-infected and CMV-free subjects. Nevertheless, a numerical simulation indicated that while this scenario might increase the risk of Type 2 error (failing to identify weak effects), it would not elevate the risk of Type 1 error (identifying non-existent effects) [77].

The fundamental experimental design of our study was cross-sectional, presenting challenges in distinguishing between causes and effects. However, our analysis went beyond merely assessing the impact of seropositivity for *Toxoplasma* or CMV. We also explored the correlation between the concentrations of specific anti-*Toxoplasma* and anti-CMV IgG antibodies. In the case of toxoplasmosis, these antibody concentrations serve as a statistical proxy for the time elapsed since infection, as demonstrated by [47]. Such correlations, combined with laboratory infection results in rodents for *Toxoplasma*, strongly suggest that the infection is likely the cause, rather than the effect, of the observed differences in cognitive performance [78]. An alternative explanation could involve an unknown third factor, such as socioeconomic status, correlating with the likelihood of CMV or *Toxoplasma* infection and intelligence. While the Czech population, particularly students of the Faculty of Science, Charles University, exhibits a relatively low level of socioeconomic stratification, future studies should consider a broader range of potential confounding variables.

Some significant effects observed in the total population (unstratified by Rh factor and sex) may seem small, yet many effects were surprisingly large for the field of biological psychology. For instance, a Tau value of 0.161 for the association between toxoplasmosis and general intelligence in men corresponds to a Cohen’s f of 0.26, considered a medium effect [79]. Similarly, a Tau of −0.32 for the association between CMV and general intelligence in Rh-negative men corresponds to a Cohen’s f of 0.55, typically deemed a large effect.

Given the exploratory nature of the main part of this study, we discussed not only formally significant effects but also nonsignificant trends and effects that lost significance after correcting for multiple tests. Confirming these effects in larger cross-sectional studies or in case-control studies enriched with Rh-negative *Toxoplasma*-infected individuals is essential.

While this sample from the Faculty of Science at Charles University, a prestigious institution, may offer insights into its student population, it’s crucial to acknowledge that these students, with an average IQ of 120, might not accurately represent the broader Czech population. Consequently, extrapolating the results of our study to a wider context should be approached with caution.

## 5. Conclusions

We have substantiated that *Toxoplasma* infection exerts numerous effects on human cognitive performance, many of which are dependent on sex and Rh phenotype. Similarly, we have extended these findings to another pathogen, the human cytomegalovirus (CMV). Given the high prevalence of latent toxoplasmosis (about 33% worldwide) and CMV infection (likely over 80% in adult populations), the observed association, such as a 2.3 IQ point decrease in *Toxoplasma*-infected individuals compared to uninfected ones (with a Cohen’s d of 0.213, categorized as a small effect), could have significant implications for the quality of life of a large number of people. These differences, observed as a decrease in IQ among infected individuals, may not only have implications for those directly affected but could also indirectly influence the broader uninfected population through societal and communal interactions.

Currently, there are no available treatments for latent toxoplasmosis or CMV infection. However, the development of vaccines against these infections could offer a preventative solution. Specifically for toxoplasmosis control, vaccinating both pet and feral cats with oral vaccines could be a pivotal strategy in eradicating the disease. Our study highlights the importance of continued research into pathogens that may seem harmless but could have significant effects on human health and wellbeing.

## Data Availability

The complete dataset is available at Figshare: https://doi.org/10.6084/m9.figshare.21865677.

## Supplementary Materials

**Supplementary Table S1.** Associations of sex, toxoplasmosis, CMV infection, and age with variables related to cognitive performance.

|  | Sex |  | *Toxoplasma* |  | CMV |  |
| --- | --- | --- | --- | --- | --- | --- |
|  | p | Cohen's d | p | Cohen's d | p | Cohen's d |
| **I-S-T 2000 R (intelligence and knowledge: N = 533, memory: N = 305)** |  |  |  |  |  |  |
| General intelligence | **0.001** | 0.307 | 0.053 | -0.213 | 0.744 | 0.029 |
| General knowledge | **0.000** | 0.731 | 0.636 | -0.051 | 0.760 | -0.027 |
| Verbal intelligence | **0.043** | 0.187 | 0.640 | -0.055 | **0.045** | 0.178 |
| Verbal knowledge | **0.000** | 0.584 | 0.640 | 0.052 | 0.151 | 0.127 |
| Numerical intelligence | **0.000** | 0.430 | 0.091 | -0.195 | 0.792 | -0.023 |
| Numerical knowledge | **0.000** | 0.441 | 0.087 | -0.186 | 0.206 | -0.112 |
| Figural intelligence | 0.577 | 0.051 | 0.096 | -0.184 | 0.794 | -0.023 |
| Figural knowledge | **0.000** | 0.692 | 0.729 | 0.037 | 0.667 | -0.038 |
| Crystallized intelligence | **0.000** | 0.725 | 0.954 | 0.006 | 0.960 | 0.004 |
| Fluid intelligence | **0.017** | 0.220 | 0.070 | -0.208 | 0.882 | 0.013 |
| Associative memory | **0.000** | -0.426 | 0.841 | -0.025 | 0.121 | -0.181 |
| **Meili memory tests (location: N = 457, free recall: N = 454, yes-no: N = 456)** |  |  |  |  |  |  |
| Spatial memory | **0.000** | -0.426 | 0.339 | -0.121 | 0.198 | -0.125 |
| Free recall memory | 0.688 | -0.038 | **0.037** | 0.203 | 0.840 | -0.020 |
| Recognition memory | **0.048** | -0.194 | 0.315 | -0.131 | 0.284 | -0.104 |
| **Simple reaction time test (N = 466)** |  |  |  |  |  |  |
| Reaction time | **0.018** | -0.230 | 0.399 | 0.118 | 0.200 | 0.123 |
| **Prepulse tests (acoustic: N = 415, visual: 420)** |  |  |  |  |  |  |
| Acoustic test, prepulse | 0.555 | -0.061 | 0.145 | 0.186 | 0.611 | 0.051 |
| Acoustic test, no prepulse | 0.728 | -0.034 | 0.259 | 0.136 | 0.969 | -0.004 |
| Visual test, prepulse | 0.443 | -0.078 | 0.995 | -0.001 | **0.021** | 0.230 |
| Visual test, no prepulse | 0.122 | -0.160 | 0.821 | 0.031 | 0.060 | 0.188 |
| **Stroop tests (test A: N = 419, test B: N = 407)** |  |  |  |  |  |  |
| Test A, correct no-reactions | 0.391 | -0.092 | 0.381 | 0.114 | 0.490 | 0.069 |
| Test A non-correct no-reactions | **0.026** | 0.289 | 0.603 | 0.068 | 0.451 | 0.075 |
| Test A reaction time | **0.026** | 0.243 | **0.001** | 0.509 | **0.014** | 0.246 |
| Test B correct no-reactions | 0.560 | 0.062 | 0.288 | 0.141 | 0.738 | 0.034 |
| Test B non-correct no-reactions | 0.367 | 0.106 | 0.232 | 0.276 | 0.237 | -0.124 |
| Test B reaction time | **0.000** | 0.464 | **0.003** | 0.460 | **0.037** | 0.213 |

*The table shows the effect size (Cohen's d) and significance of t-test (p). Statistically significant effects are highlighted in bold.*

## Author Contributions

Conceptualization, J.F.; Methodology, J.F. and P.K.; Validation, Š.K.; Formal Analysis, J.F.; Investigation, V.C., L.P., P.T., B.Š, Š.K.; Data Curation, J.F. and B.Š; Writing – Original Draft Preparation, J.F.; Writing – Review & Editing, J.F., V.C., L.P., P.T., B.Š and Š.K.; Supervision, J.F..; Project Administration, J.F.; Funding Acquisition, J.F.

## Institutional Review Board Statement

The study was conducted according to the guidelines of the Declaration of Helsinki, and approved by the Institutional Review Board (or Ethics Committee) of IRB of the Faculty of Science, Charles University (protocol codes 2009/06, and 2013/07, dates 10.8. 2009 and 10.2. 2013).

## Data Availability Statement

The complete dataset is available at Figshare: https://doi.org/10.6084/m9.figshare.21865677.

## Acknowledgments

We want to thank Anna Pilátová for proofreading and editing.

